# Impact of climate on COVID-19 transmission: A case study with Indian states

**DOI:** 10.1101/2020.07.05.20146324

**Authors:** Souvik Manik, Manoj Mandal, Sabyasachi Pal, Subhradeep Patra, Suman Acharya

## Abstract

Coronavirus Disease 2019 (COVID-19) started in Wuhan province of China in November 2019 and within a short time, it was declared as a worldwide pandemic by the World Health Organisation due to the very fast worldwide spread of the virus. There were a few studies that look for the correlation with infected individuals and different environmental parameters using early data of COVID-19 but there was no study that deal with the variation of effective reproduction number and environmental factors. Effective reproduction number is the driving parameter of the spread of a pandemic and it is important to study the effect of various environmental factors on effective reproduction numbers to understand the effect of those factors on the spread of the virus. We have used time-dependent models to investigate the variation of different time-dependent driving parameters of COVID-19 like effective reproduction number and contact rate using data from India as a test case. India is a large population country that was highly affected due to the COVID-19 pandemic and has a wide span of different temperature and humidity regions and is ideal for such study. We have studied the impact of temperature and humidity on the spread of the virus of different Indian states using time-dependent epidemiological models SIRD, and SEIRD for a long time scale. We used a linear regression method to look for any dependency between the effective reproduction number with the relative humidity, absolute humidity, and temperature. The effective reproduction number showed a negative correlation with both relative and absolute humidity for most of the Indian states, which are statistically significant. This implies that relative and absolute humidity may have an important role in the variation of effective reproduction numbers. There was no conclusive evidence of a correlation between effective reproduction numbers and average air temperature.

## 1 Introduction

The first case of COVID-19 was reported in India on 30 January 2020 [1]. Till 10 October 2021, India had 33.9 million total confirmed cases which were highest in Asia [2] and the second-highest in the world [2]. During the above-mentioned period, India had 33.2 million recoveries, and 0.45 million was deceased [2]. Study with India is interesting as it is the world’s second-largest population with about one-sixth of the world’s total population [3].

COVID-19’s second wave started in India on February 11, 2021. The second wave had a significant impact, with daily cases nearly four times than during the first wave’s peak [4]. In India, the second wave was caused mainly by the double mutant variant.

Currently, four mutant Coronavirus variants are dominant: a) Brazil variant [P.1 (P.1)] [5], b) Indian variant [B.1.617], c) South Africa variant [501Y.V2 (B.1.351)], and d) UK Variant [VOC 202012/01 (B.1.1.7)] [6]. WHO cautioned that the Indian variety, B.1.617, was rapidly spreading in India, accounting for more than 28% of positive test samples. The Indian strain (B.1.617) of SARS-CoV2 was found to be more infectious compared to other SARS-CoV2 variants [7].

The spread of an epidemic depends on a large number of factors. Earlier, it was found that humidity is one of the driving factors to explain the seasonality of influenza [8]. Similar seasonal variability would be expected for COVID-19 and other coronaviruses like SARS. The spread of COVID-19 (SARS-CoV-2) was found to be correlated with temperature and latitude [9]. A study of the seasonality of many other viruses, like seasonal Influenza, suggests that the cool season in the northern hemisphere favors disease propagation more than the warm season in the northern hemisphere [9]. Earlier, it was shown that 50% of the influenza virus transmission variability can be explained by absolute humidity but a detailed study was not possible for limited data [8]. Meteorological parameters (temperature, relative humidity, and absolute humidity) may play an important role in influencing infectious diseases such as severe acute respiratory syndrome (SARS) and influenza.

At the beginning of the spread of the COVID-19 disease, it was widely believed that summer will cause the virus to reduce [10]. Results of other coronaviruses (such as SARS coronavirus) also indicated that the virus may be less effective with rising temperature and humidity [11]. A rigorous study was necessary with a large amount of data to explore the effect of temperature and humidity on the spread of COVID-19.

There were several social or non-meteorological factors that could influence the correlation study between meteorological factors and COVID-19 transmissions, such as governmental interventions, social interactions, lock-downs, herd immunity, vaccinations, population density, personal hygiene, defense mechanisms, and cultural activities. Several studies focused on the impact of social interaction on COVID-19 spread. For example, it was found that the 2021 assembly election played a major role in the COVID-19 spread rate in India [12].

In the present paper, we investigate the influence of average air temperature, absolute humidity, and relative humidity on the effective contact rate and reproduction number of COVID-19. There were a few studies that look for the correlation with infected individuals and different environmental parameters using early data of COVID-19 but so far there was no study which deals with the variation of effective reproduction number and environmental factors. It is crucial to study different time-dependent models to account for the real scenario of a pandemic. Effective reproduction number is the driving parameter of the spread of a pandemic and it is important to study the effect of various environmental factors on effective reproduction number to understand the effect of those factors on the spread of the virus. So, we have done a rigorous study for different Indian states which were badly affected during the second wave of COVID-19. India also represented a wide variety of temperatures and humidity regions, which was ideal for our study. In this paper, we have looked at the variety of different driving parameters of the pandemic like effective contact rate (*β*_*n*_) and effective reproduction number (ℝ(t)) using time-dependent mathematical models SIRD and SEIRD for different Indian states. One of the important goals of this study is to investigate the impact of temperature and humidity on effective reproduction number.

In Sect. 2, we summarised different mathematical models of the pandemic. Sect. 2.1 carried a brief introduction of the SIRD model. A short introduction of the SEIRD model was given in Sect. 2.2. In Sect. 4, we presented our findings. The temperature dependence of the effective reproduction number was discussed in the Sect. 4.1 and the humidity dependence of effective reproduction number ℝ(*t*) was discussed in Sect. 4.2. In Sect. 5, we discussed our findings, and in Sect. 6 we summarised our results.

## 2 Mathematical models

Different mathematical models were designed to simulate the effect of the disease from different perspectives [13, 14, 15, 16, 17]. These helped us to get an idea about the spread rate of a disease over the population. We have used the most efficient time-dependent epidemiological models SIRD and SEIRD. The spread rate depends on a large number of parameters such as mean incubation time [18, 19], mean infectious period, social distancing, and risk of international spread [20, 21]. The most important part of these models is to calculate the basic reproduction number (*R*_0_) which is the contagiousness of the diseases. *R*_0_ is defined as the average number of people who can be affected by a single infected person over a course of time. *R*_0_ > 1 indicates that the spread is increasing, *R*_0_ = 1, indicates that the spread is stable and *R*_0_ < 1, indicates that the spread is expected to stop. We studied the effective reproduction number ℝ(*t*) for India using different models. We also studied the evolution of ℝ(*t*) using the same models for different individual states of India.

### 2.1 SIRD model

There are several models for studying contagious disease transmission in a large population. The simplest compartmental model that can explain the evolution of an outbreak at the population level is the Susceptible-Infected-Recovered (SIR) model. The SIRD model [16, 12] is the extended version of the SIR model. We used this model to study different time-dependent parameters like effective reproduction number (ℝ), contact rate (*β*), recovery rate (*γ*), and mortality rate (*δ*). At any time *t, S*(*t*) be the total number of susceptible individuals, *I*(*t*) be the total number of infected individuals, *R*(*t*) be the total number of recovered, and *D*(*t*) be the total number of deceased individuals.

The infant entered susceptible class *S* once the maternal antibodies no longer existed in the body. Since a susceptible has enough contact with an infective to allow transmission, they are known as infective class *I*, which means they are contagious in the case they can transmit the virus. Once the infectious phase is elapsed, the individual falls into one of two classes: *R*, which is made up of people who gained immunity through infection, or *D*, which is made up of deceased people. Here, *β* is the contact rate and *γ* is the recovery rate and *δ* is the mortality rate. *s*(*t*), *i*(*t*), *r*(*t*) and *d*(*t*) can be expressed in fractional form:

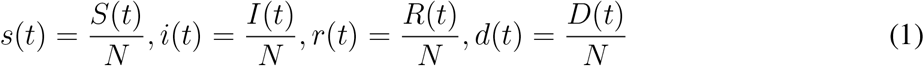

from the conservation law,

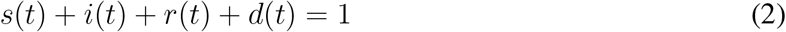

SIRD model can be expressed by following set of differential equations.

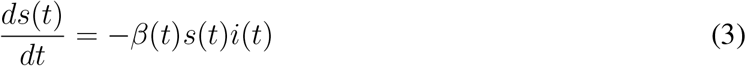

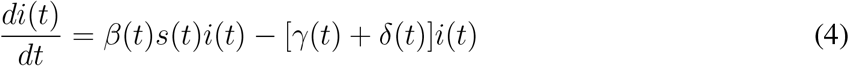

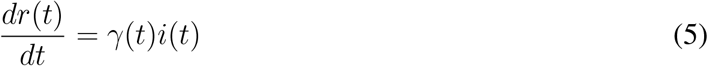

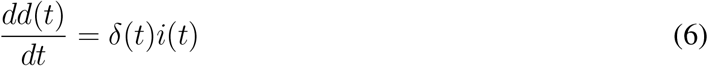

from the equation 4, we may write

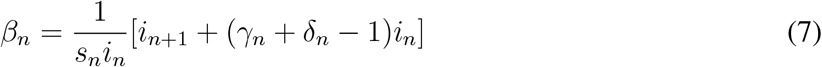

We use equation (5) to find *γ*_*n*_

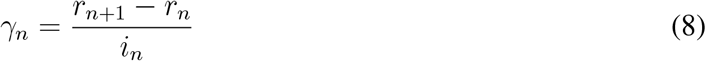

Now we use equation (6) to find *δ*_*n*_

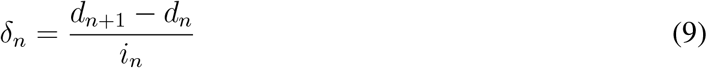

The effective reproduction number R(t) can be expressed as

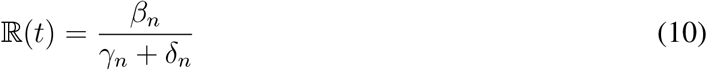

In Fig. 3, we have shown a variety of different parameters for the SIRD model for few selected Indian states. In the second, third, and fourth row, the variation of effective contact rate (*β*_SIRD_(*t*)), the variation of recovery rate (*γ*_SIRD_(*t*)), and the variation of mortality rate(*δ*_SIRD_(*t*)) were shown. The fifth row represents the variation of effective reproduction number (ℝ _SIRD_(*t*)) for the SIRD model.

### 2.2 SEIRD model

The Susceptible-Exposed-Infected-Recovered-Deceased (SEIRD) model [22] is the extended version of the SEIR model. At any time *t*, *S*(*t*) be the total number of susceptible individuals, *E*(*t*) be the total number of exposed individuals, *I*(*t*) be the total number of infected individuals, *R*(*t*) be the total number of recovered individuals, and *D*(*t*) be the total number of deceased individuals from the epidemic. In the SEIRD model, we considered the average incubation period of 1*/σ* as a constant. We presume that the individuals who recovered from the disease can not become susceptible again.

Various compartments of the SEIRD model were presented in Fig. 2. The Exposed (*E*) compartment in the SEIRD model is an extension to the SIRD model. The susceptible (*S*) moves into the exposed compartment (*E*) when a susceptible individual comes across significant interaction with an infective individual (*I*). The individual entered the class *I* at the end of the latent phase and became capable to spread the infection. Finally, at the end of the infectious time, the person moved to either the recovered compartment *R* or the deceased compartment *D* (for detail, see section 2 of [12]).

The parameter *β* is the product of the average number of contacts per person and per unit time by the probability of disease transmission in contact between susceptible and infectious individuals. *γ* is the recovery rate. *δ* is the mortality rate. The compartment *E* of the exposed individuals in the SEIRD model makes the model slightly more delicate. The SEIRD model is a compartmental model used to understand the mathematical modeling of infectious diseases in a large population. If *N* is the total population size then in fraction form

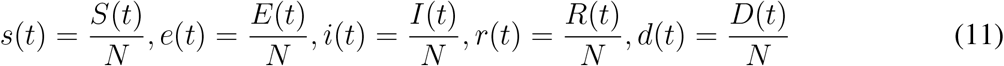

From the conservation of the total number of individuals, we have the relation

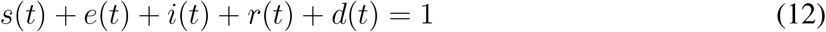

These quantities change by discrete amounts but if the minimal possible change is relatively low, we can consider *s*(*t*), *i*(*t*), *r*(*t*), *d*(*t*) as differentiable functions. For any time *t* ≥ 0, the SEIRD model can be expressed by the differential equations

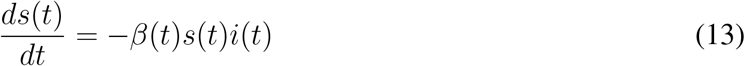

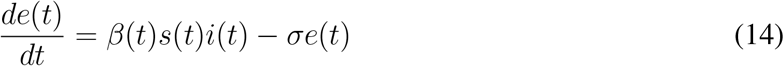

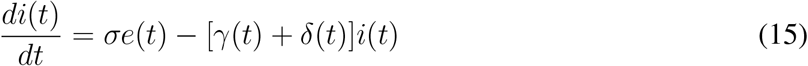

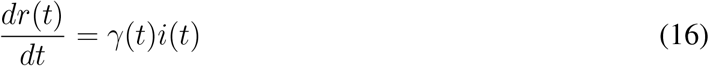

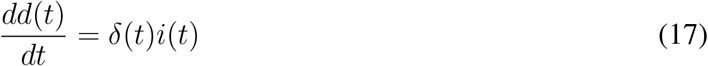

We use the discrete form of equations (13) to (17).

We use equation (14) to find *β*_*n*_

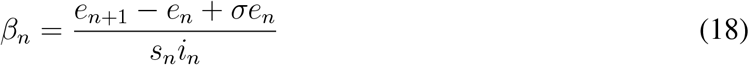

We use equation (16) to find *γ*_*n*_

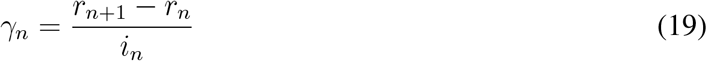

Now we use equation (17) to find *δ*_*n*_

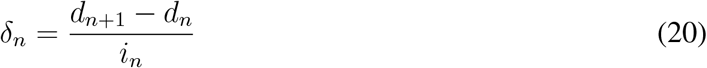

The effective reproduction number ℝ(t) can be given by

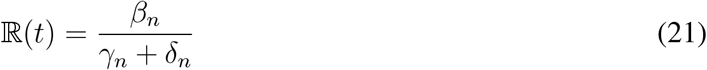

In Fig. 3, we have shown the variation of different parameters for the SEIRD model for a few selected Indian states. In the second and third row, the variation of effective contact rate (*β*_SEIRD_(*t*)) and the variation of recovery rate (*γ*_SEIRD_(*t*)) were shown. The fourth row showed the variation of mortality rate (*δ*_SEIRD_(*t*)) and the fifth row showed the variation of effective reproduction number (ℝ_SEIRD_(*t*)) for the SEIRD model.

## 3 Data Analysis and Methodology

We used the COVID-19 data maintained by the Center for Systems Science and Engineering (CSSE) at Johns Hopkins University to track reported cases of Coronavirus in real-time [23]. The time-series data of different Indian states can be accessed from these source^1^. We obtained weather data from “World Weather Online” using a python based API from 1st February to 10th October 2021. We taken state population data from “India Census” site^2^. We have used standard python packages i.e., Numpy, Scipy, Pandas, Matplotlib for the entire work. Since environmental variables were not expected to have an immediate effect on COVID-19 cases, we acquired weather data including temperature and relative humidity 7 days before the reported cases. We also calculated absolute humidity, the actual amount of water vapor in the atmosphere, depending on the air temperature (*T*; in *°C*), and relative humidity (RH; in %) [24]. The absolute humidity (AH; in *g/m*^3^) is the weight of water vapor per unit volume of air and was estimated using the Clausius–Clapeyron equation and can be described as follows [25].

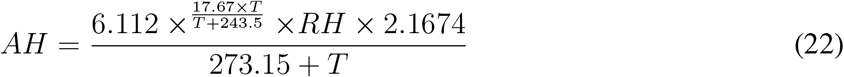

where *AH* corresponds to absolute humidity, *RH* is the relative humidity, and *T* is the average temperature.

We have selected the ten most affected Indian states for which the total confirmed COVID-19 cases were maximum during the time-span of COVID-19 second wave in India (2 February 2021 to 10 October 2021). The lower threshold of total confirmed individuals was 0.56 million during the abovementioned period for those selected states. We studied the variation of confirmed, recovered, and deceased individuals for Indian states Maharashtra, Kerala, Karnataka, Tamil Nadu, Andhra Pradesh, Uttar Pradesh, West Bengal, Delhi, Rajasthan, and Gujarat. These states are located at different corners of the country with a wide variability of different atmospheric parameters. The effective contact rate (*β*(*t*)), recovery rate (*γ*(*t*)), mortality rate (*δ*(*t*)) and effective reproduction number (ℝ(*t*)) were estimated from the mathematical equations discussed in Sect. 2 for different states of India. We also studied the evolution of these parameters using times series data. Since some of the states have wide variability of different atmospheric parameters, we have taken the averaged value of different meteorological data across different cities for every state.

We calculated the values of transmission coefficients using SIRD and SEIRD models (for details, Sect. 2.1, 2.2) for different states (Maharashtra, Kerala, Karnataka, Tamil Nadu, Andhra Pradesh, Uttar Pradesh, West Bengal, Delhi, Rajasthan, and Gujarat) of India. To minimize the effect of noise, we used a three-day rolling mean variation of transmission coefficients for our study. For the SIRD model, we used equation (7), (8), (9) and (10) to calculate the effective contact rate (*β*(*t*)), recovery rate (*γ*(*t*)), mortality rate (*δ*(*t*)) and effective reproduction number (ℝ(*t*)) respectively. For the SEIRD model, we used equation (18), (19), (20) and (21) to calculate *β*(*t*), *γ*(*t*), *δ*(*t*), and ℝ(*t*) respectively. We optimized the values of all transmission coefficients by removing all negative values. Linear regression was carried out between the 7-days rolling mean of the various meteorological variables and the effective reproduction number (ℝ(*t*)) to understand the effect of meteorological parameters on the spread of COVID-19.

## 4 Result

For our study, we used the time series data for infected, recovered, and deceased individuals of COVID-19 [23]. The impact of temperature and humidity on ℝ(*t*), *β*(*t*), and *γ*(*t*) was studied in detail for a large population, which is very relevant to the practical scenario. In the first row of Fig. 1, we showed the daily variation of the number of confirmed individuals for India. In the second row of Fig. 1, we showed the variation of the deceased individuals. From the figure, two different waves of the COVID-19 in India were visible. The second wave was nearly four times the first wave and gave ideal time to investigate the correlation of the spread rate of the virus with environmental parameters for a large population. The shaded region showed the duration of our study.

**Figure 1:**
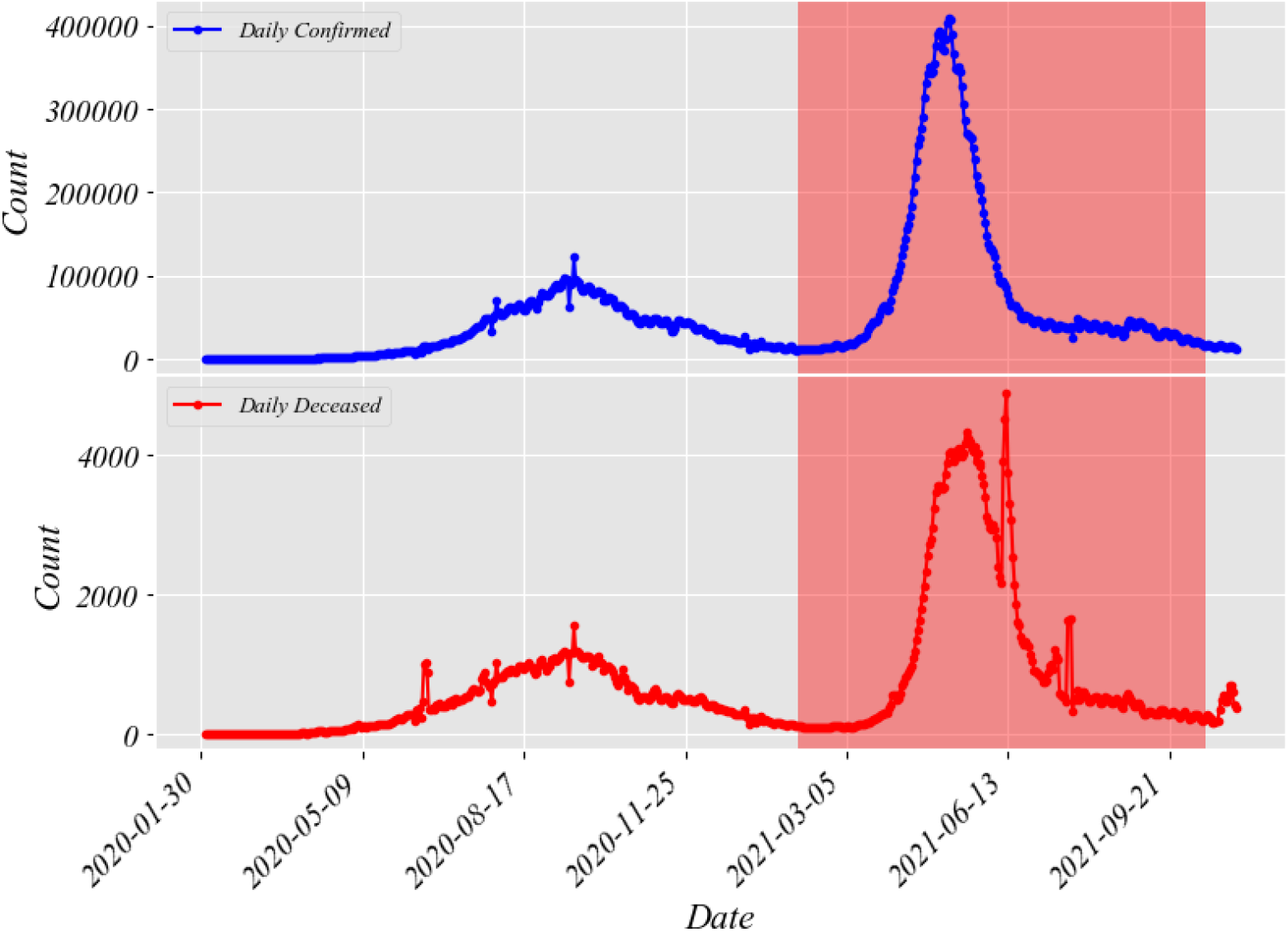
Variation of daily confirmed and deceased individuals for India with 3 days rolling mean. The shaded region showed the time for which the whole study was conducted during the second wave of COVID-19 in India.

In Fig. 3, we showed examples of the variation of different parameters for eight selected Indian states which were badly affected by the coronavirus. We showed the variation of driving parameters of a pandemic for the states Maharashtra, Kerala, Karnataka, Tamil Nadu, Andhra Pradesh, Uttar Pradesh, West Bengal, Delhi, Rajasthan, and Gujarat. These states belong to different corners of the country with wide variability for different meteorological parameters like temperature and humidity and formed an ideal sample of states to study the impact of temperature and humidity on the spread of the virus. In Fig. 3, the first row showed the variation of the number of infected, recovered, and deceased individuals with time, the second row showed the variation of effective contact rate *β*(*t*) with time, the third row showed the variation of recovery rate *γ*(*t*) with time, the fourth row showed the variation of deceased individuals and the fifth row showed the variation of effective reproduction number ℝ(*t*) with time for different Indian states. Fig. 4 showed the variation of average temperature *T*_*av*_, relative humidity *RH* and absolute humidity *AH* for different Indian states for which the study is executed. The figure showed a wide range of temperature and humidity since selected states represented different regions of the country.

**Figure 2:**
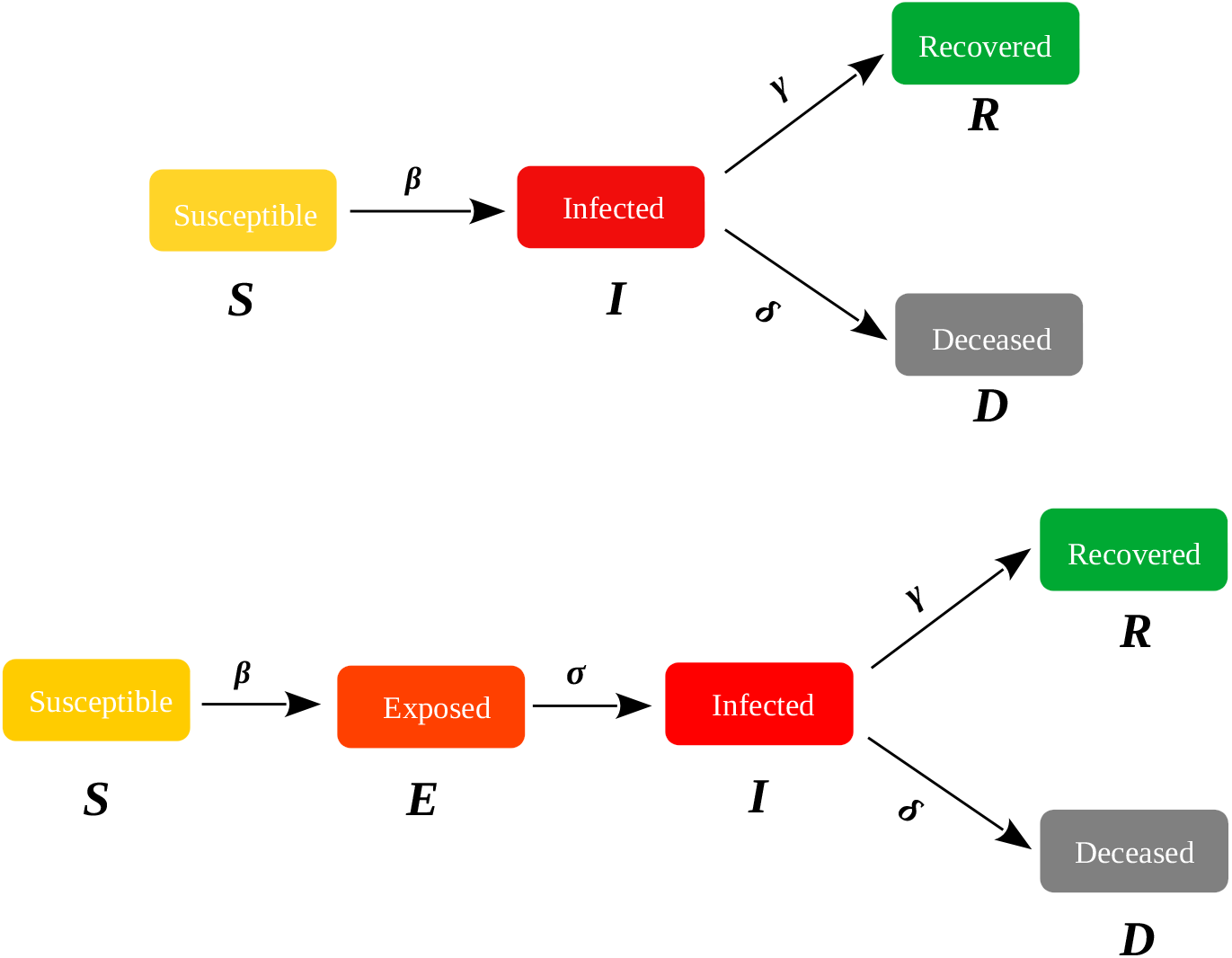
The schematic diagram showing different compartments of SIRD, and SEIRD model. *β* is the contact rate, *γ* is the recovery rate, *σ* is the incubation period, and *δ* is the mortality rate.

**Figure 3:**
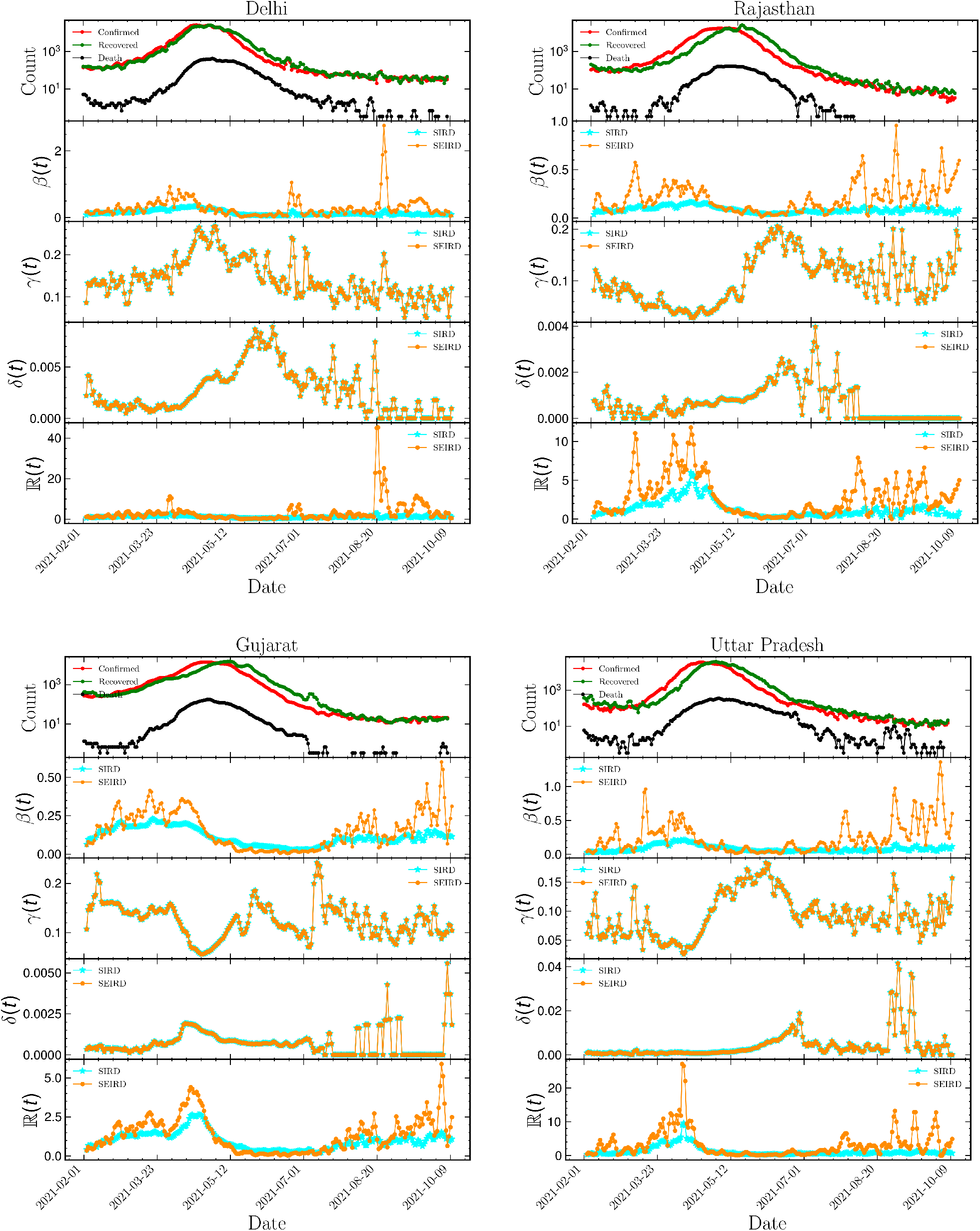
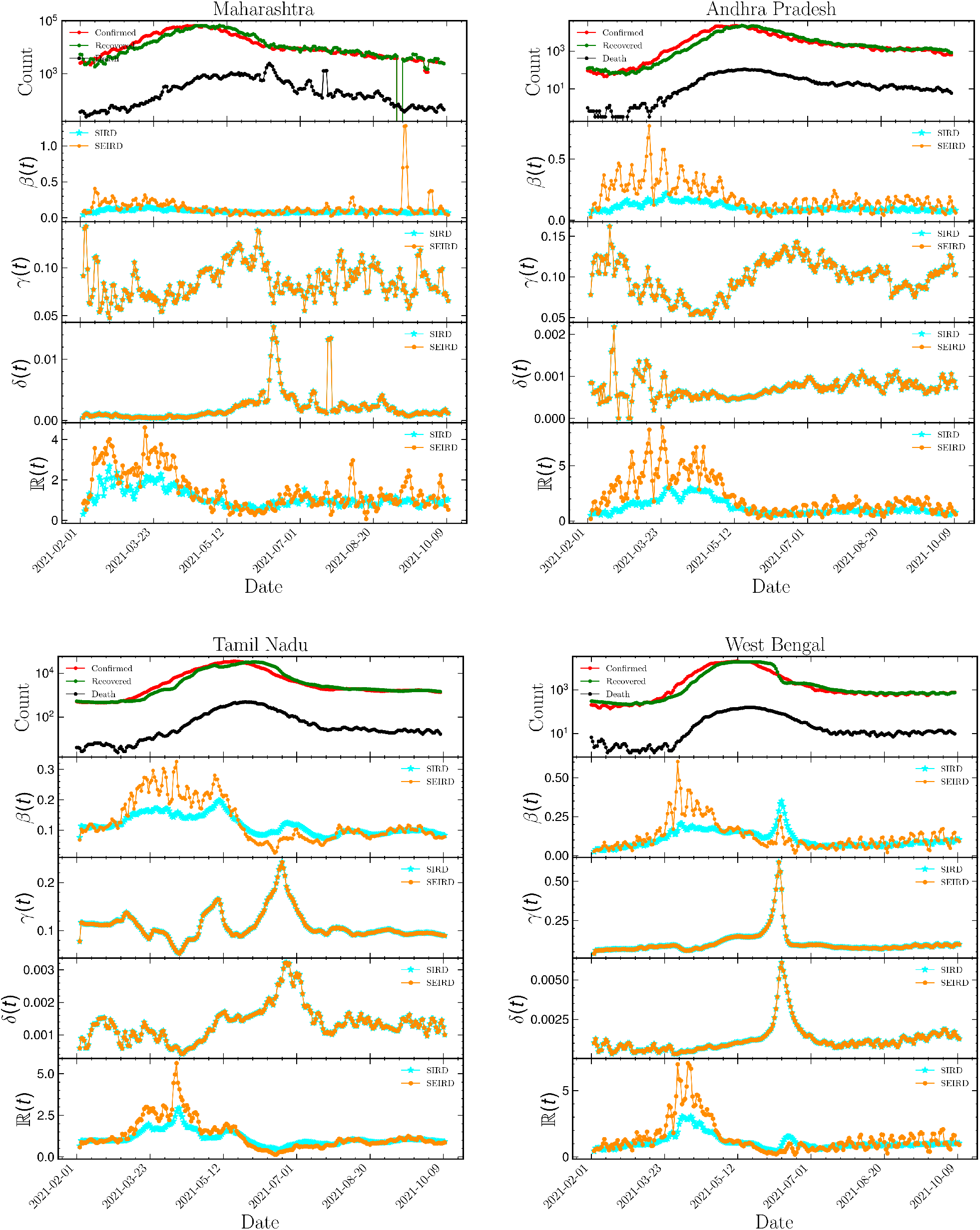
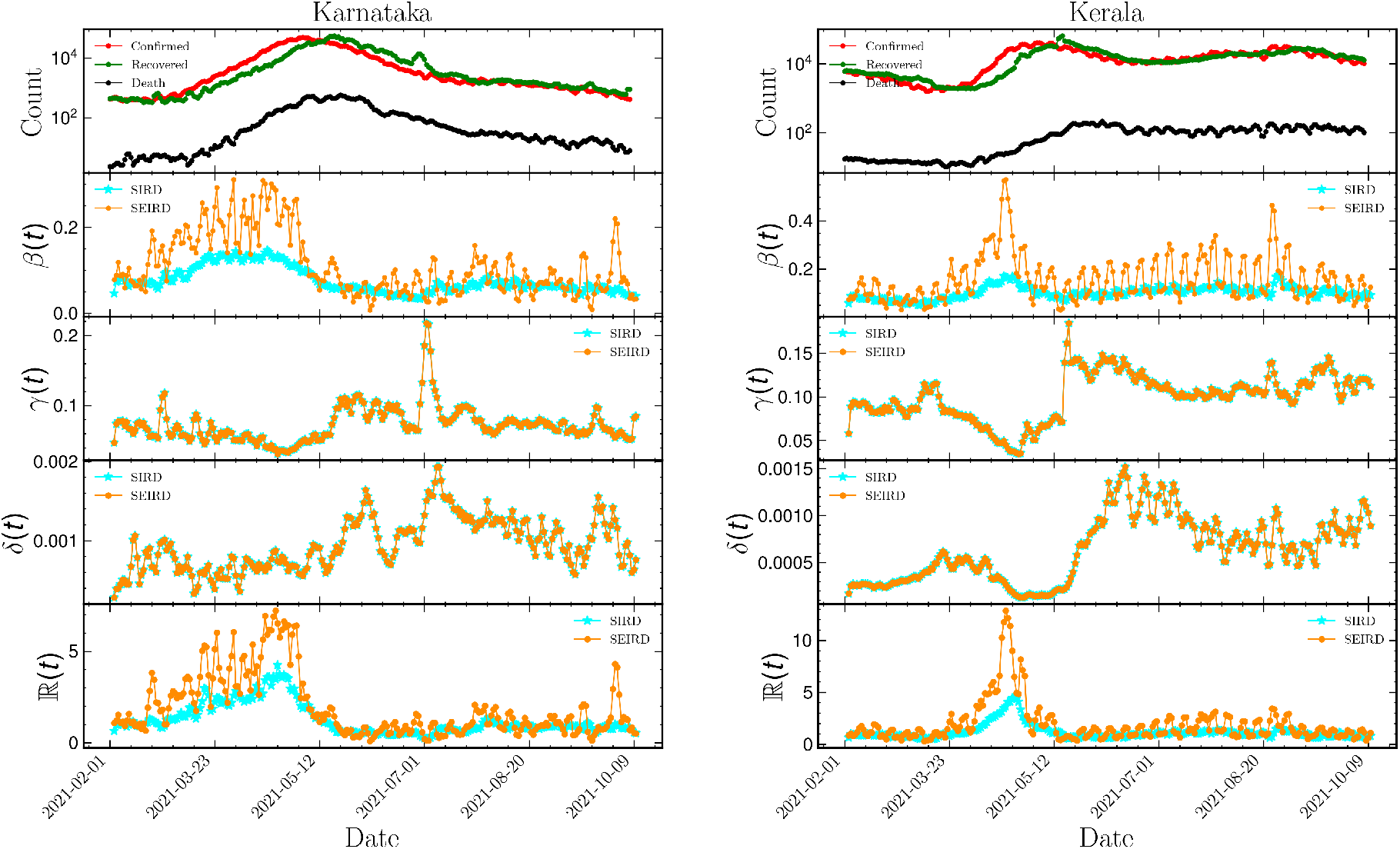
Variation of different driving parameters of the pandemic with time for different Indian states. *γ*(*t*) is the recovery rate, *β*(*t*) is the effective contact rate, *δ*(*t*) is the mortality rate and ℝ(*t*) is the effective reproduction number.

**Figure 4:**
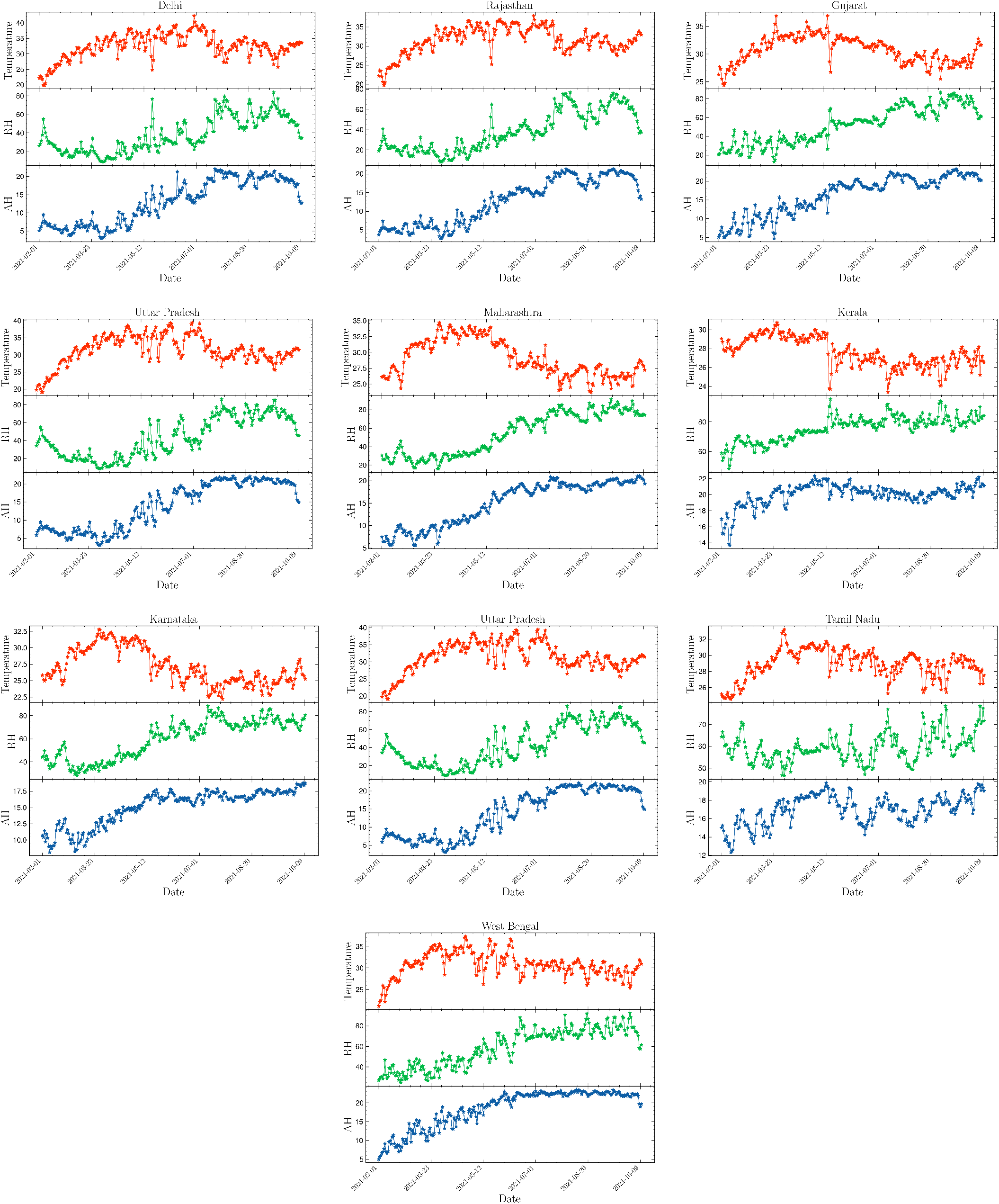
Variation of average temperature, relative humidity (RH) and absolute humidity (AH) with time for different Indian states.

### 4.1 Temperature dependence of ℝ(*t*)

We studied the dependence of effective reproduction number ℝ(*t*) on temperature for different Indian states. The linear regression method was used to find the correlation and the regression coefficient *R*^2^. Table 1 showed the result of the linear regression method for different models for the study period. The positive sign indicated the correlation between effective reproduction number (ℝ(*t*)) and average temperature (*T*_*av*_) with a positive slope and the negative sign indicated a correlation between ℝ(*t*) and temperature with a negative slope. The correlation with negative slope indicated that the effective reproduction number (ℝ(*t*)) will decrease with the increase of the temperature. For the SIRD model, two states Delhi and Rajasthan showed a negative correlation between ℝ(*t*) and *T*_*av*_. Eight states showed a positive correlation between ℝ(*t*) and *T*_*av*_.

**Table 1:**
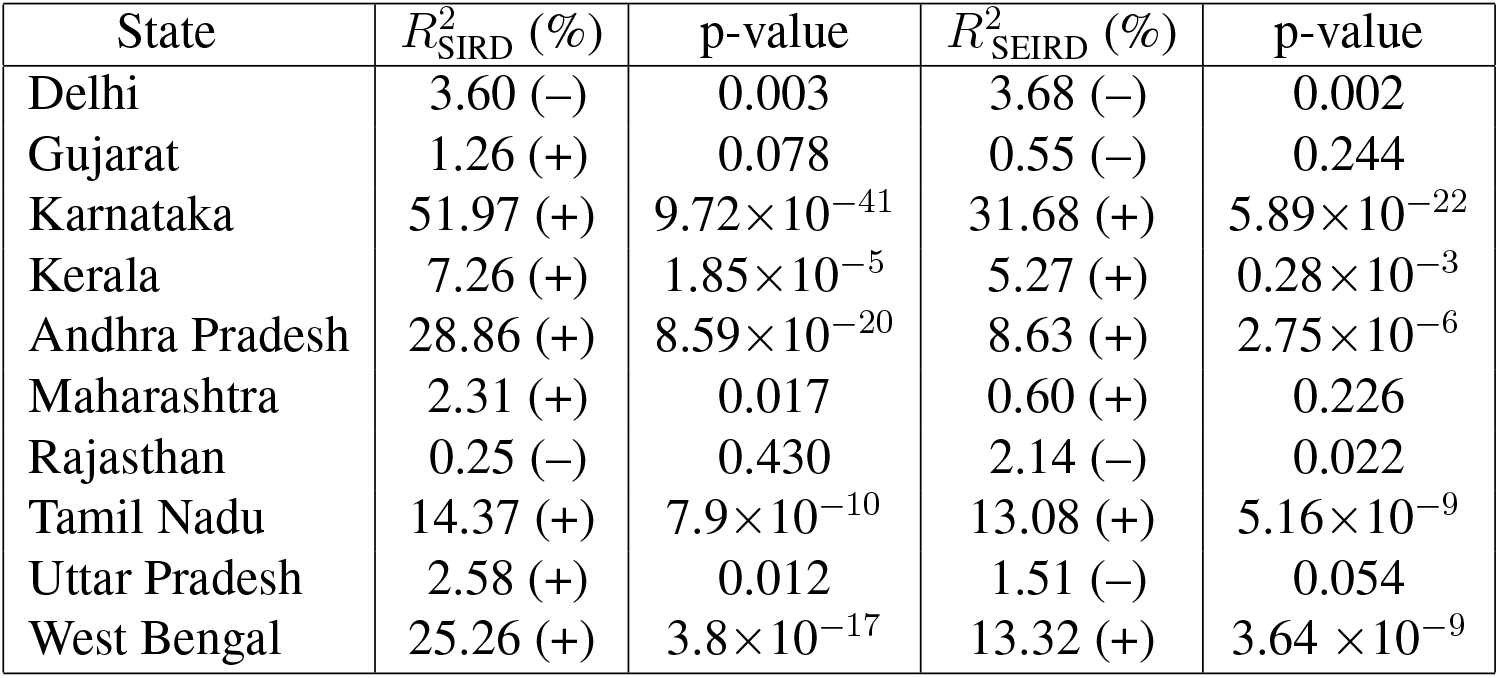
Value of regression coefficient *R*^2^ with p-value using different models for different Indian states from ℝ(t) vs *T*_*av*_ plot with seven days running average. The ‘+’ sign implies positive correlation and ‘–’ sign implies negative correlation between ℝ(t) and *T*_*av*_.

| State | $R^2_{\text{SIRD}} (\%)$ | p-value | $R^2_{\text{SEIRD}} (\%)$ | p-value |
| --- | --- | --- | --- | --- |
| Delhi | 3.60 (–) | 0.003 | 3.68 (–) | 0.002 |
| Gujarat | 1.26 (+) | 0.078 | 0.55 (–) | 0.244 |
| Karnataka | 51.97 (+) | $9.72 \times 10^{-41}$ | 31.68 (+) | $5.89 \times 10^{-22}$ |
| Kerala | 7.26 (+) | $1.85 \times 10^{-5}$ | 5.27 (+) | $0.28 \times 10^{-3}$ |
| Andhra Pradesh | 28.86 (+) | $8.59 \times 10^{-20}$ | 8.63 (+) | $2.75 \times 10^{-6}$ |
| Maharashtra | 2.31 (+) | 0.017 | 0.60 (+) | 0.226 |
| Rajasthan | 0.25 (–) | 0.430 | 2.14 (–) | 0.022 |
| Tamil Nadu | 14.37 (+) | $7.9 \times 10^{-10}$ | 13.08 (+) | $5.16 \times 10^{-9}$ |
| Uttar Pradesh | 2.58 (+) | 0.012 | 1.51 (–) | 0.054 |
| West Bengal | 25.26 (+) | $3.8 \times 10^{-17}$ | 13.32 (+) | $3.64 \times 10^{-9}$ |

The first column of Fig. 5 represented the variation of the effective reproduction number ℝ(*t*) with average temperature *T*_*av*_ for different Indian states (Maharashtra, Kerala, Karnataka, Tamil Nadu, Andhra Pradesh, Uttar Pradesh, West Bengal, Delhi, Rajasthan, and Gujarat). The variation of ℝ(*t*) with *T*_*av*_ was estimated for two different time-dependent models SIRD and SEIRD. A positive correlation was found for the states West Bengal, Tamil Nadu, Maharastra, Andhra Pradesh, Kerala, and Karnataka for both models, whereas the states Delhi, and Rajasthan showed a negative correlation between ℝ(*t*) and *T*_*av*_. For the state of Gujarat and Uttar Pradesh, the correlation results were not conclusive. The state Gujarat showed a positive correlation with *T*_*av*_ for the SIRD model and a negative correlation for the SEIRD model, which was less significant than the SIRD model. For Uttar Pradesh, the SIRD model showed a positive correlation and the SEIRD model showed a negative correlation, both correlations were statistically less significant (p-value>0.05).

**Figure 5:**
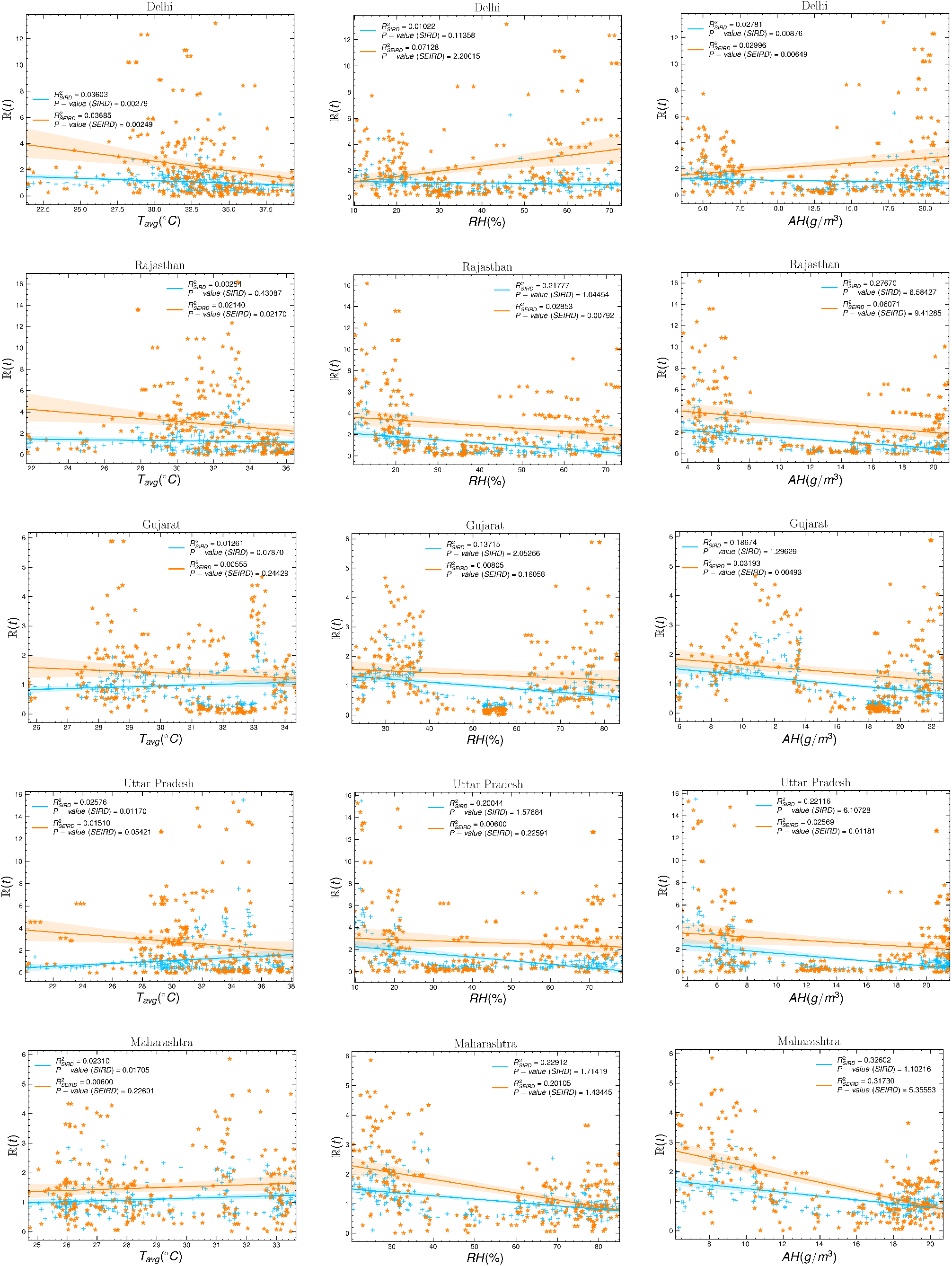
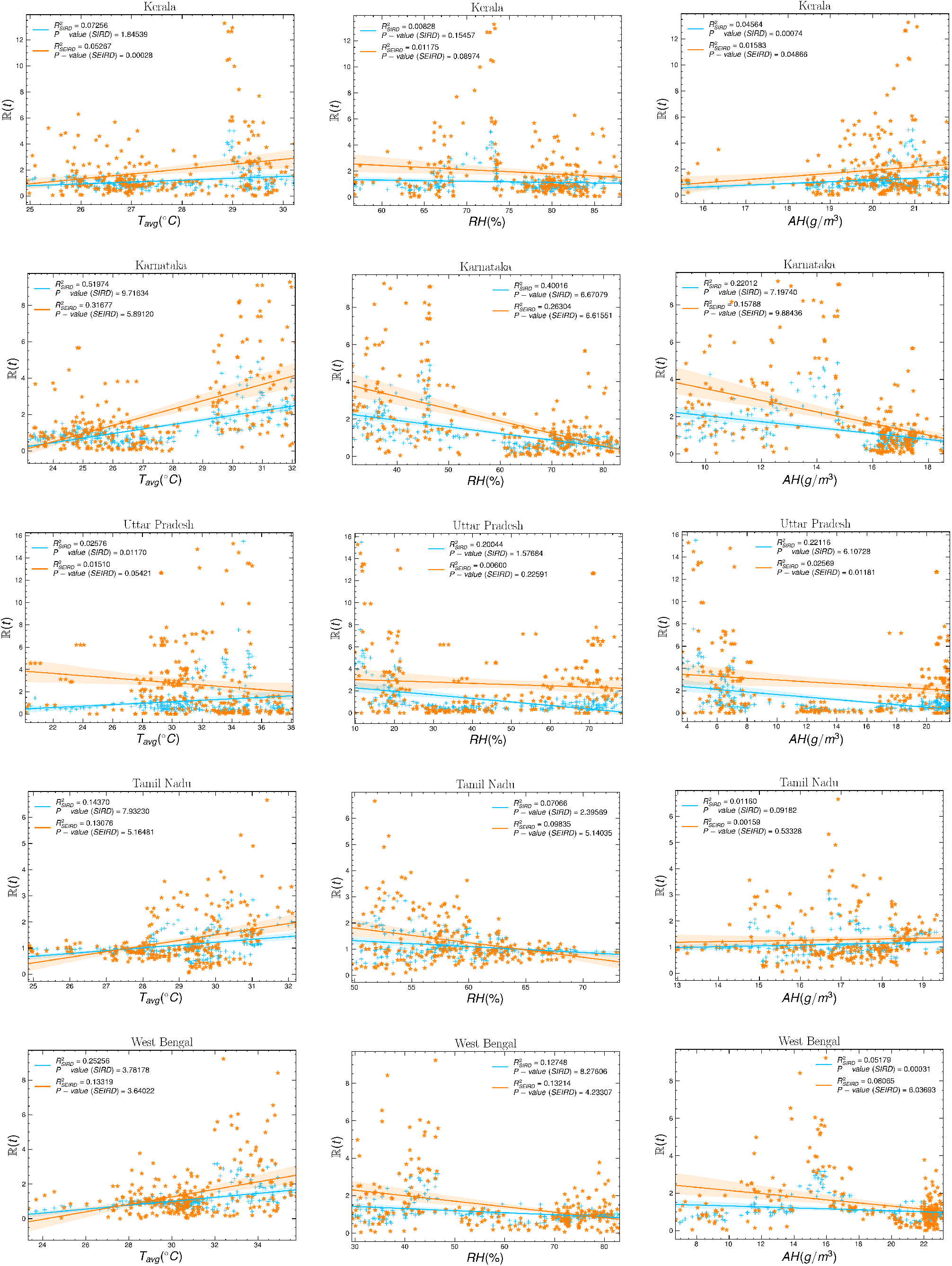
Correlation between the effective reproduction number ℝ(*t*) and 7 days moving average of the different meteorological parameters.

### 4.2 Humidity dependence of ℝ(t)

Humidity plays a crucial role as a driving environmental parameter on the spread of a virus. We investigated whether there is any dependence of relative and absolute humidity on the spread rate of COVID-19. We studied the correlation of effective reproduction number ℝ(*t*) with relative humidity (*RH*) and absolute humidity (*AH*) with a seven-day running average. We have used the linear regression method to look for any correlation between ℝ(*t*) with *RH* and *AH*. We find out the correlation coefficient *R*^2^. We summarised the result of variation of the ℝ(*t*) with average *RH* and *AH* for different states of India in Table 2. For the SIRD model, all ten states showed a negative correlation between ℝ(*t*) with *RH*. The SEIRD model also showed that all states followed a negative correlation except Delhi. Except for Delhi, the negative correlation is highly significant for both models.

**Table 2:** The summary of regression coefficient *R*^2^ with p-value using different models for different Indian states from ℝ(t) vs *RH* plot with seven days running average. The ‘+’ sign implies positive correlation and ‘–’ sign implies negative correlation between ℝ(t) and *RH*.

| State | $R^2_{\text{SIRD}}$ (%) | p-value | $R^2_{\text{SEIRD}}$ (%) | p-value |
| --- | --- | --- | --- | --- |
| Delhi | 1.02 (–) | 0.113 | 7.13 (+) | $2.20 \times 10^{-5}$ |
| Gujarat | 13.71 (–) | $2.05 \times 10^{-9}$ | 0.80 (–) | 0.160 |
| Karnataka | 40.01 (–) | $6.67 \times 10^{-29}$ | 26.30 (–) | $6.61 \times 10^{-18}$ |
| Kerala | 0.83 (–) | 0.154 | 1.17 (–) | 0.089 |
| Andhra Pradesh | 32.70 (–) | $9.25 \times 10^{-23}$ | 24.73 (–) | $8.99 \times 10^{-17}$ |
| Maharashtra | 22.91 (–) | $1.71 \times 10^{-15}$ | 20.10 (–) | $1.43 \times 10^{-13}$ |
| Rajasthan | 21.78 (–) | $1.044 \times 10^{-14}$ | 2.85 (–) | 0.008 |
| Tamil Nadu | 7.07 (–) | $2.39 \times 10^{-5}$ | 9.83 (–) | $5.14 \times 10^{-7}$ |
| Uttar Pradesh | 20.04 (–) | $1.57 \times 10^{-13}$ | 0.60 (–) | 0.225 |
| West Bengal | 12.75 (–) | $8.27 \times 10^{-9}$ | 13.21 (–) | $4.23 \times 10^{-9}$ |

The second column of Fig. 5 showed the correlation of the ℝ(*t*) with *RH* for ten Indian states which were badly affected by COVID-19. Most of the states showed a negative correlation between ℝ(*t*) and *RH*, which implied that the relative humidity might be playing a role to control the spread of the virus.

We also looked for the correlation between ℝ(*t*) and *AH*. The result of variation of ℝ(*t*) with *AH* is summarized in Table 3. For the time-dependent SIRD model, eight states out of ten states showed a negative correlation between ℝ(*t*) and *AH*, which were statistically significant (p-value<0.05). For the SEIRD model also most of the states showed a negative correlation between ℝ(*t*) and *AH*. For the state of Tamil Nadu, the correlation was positive for both the models SIRD and SEIRD but the correlation was statistically less significant (p-value>0.05). For the state of Delhi, we have not found a conclusive result, though the SEIRD model showed a positive correlation with a higher statistical significance than the SIRD model. For the state of Kerala, we found a positive correlation between ℝ(*t*) and *AH* for both the models SIRD and SEIRD.

**Table 3:**
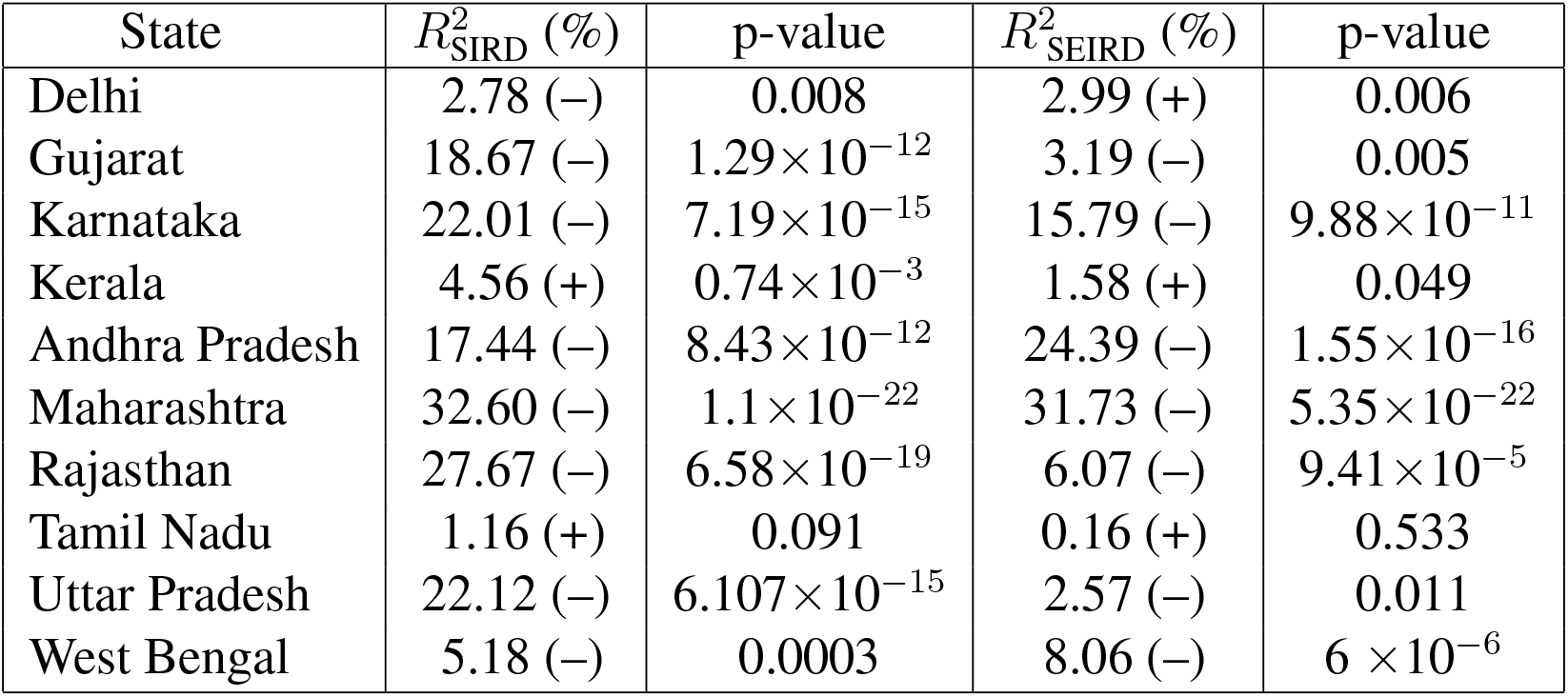
The summary of regression coefficient *R*^2^ with p-value using different models for different Indian states from ℝ(t) vs *RH* plot with seven days running average. The ‘+’ sign implies positive correlation and ‘–’ sign implies negative correlation between ℝ(t) and *AH*.

| State | $R^2_{\text{SIRD}}$ (%) | p-value | $R^2_{\text{SEIRD}}$ (%) | p-value |
| --- | --- | --- | --- | --- |
| Delhi | 2.78 (–) | 0.008 | 2.99 (+) | 0.006 |
| Gujarat | 18.67 (–) | $1.29 \times 10^{-12}$ | 3.19 (–) | 0.005 |
| Karnataka | 22.01 (–) | $7.19 \times 10^{-15}$ | 15.79 (–) | $9.88 \times 10^{-11}$ |
| Kerala | 4.56 (+) | $0.74 \times 10^{-3}$ | 1.58 (+) | 0.049 |
| Andhra Pradesh | 17.44 (–) | $8.43 \times 10^{-12}$ | 24.39 (–) | $1.55 \times 10^{-16}$ |
| Maharashtra | 32.60 (–) | $1.1 \times 10^{-22}$ | 31.73 (–) | $5.35 \times 10^{-22}$ |
| Rajasthan | 27.67 (–) | $6.58 \times 10^{-19}$ | 6.07 (–) | $9.41 \times 10^{-5}$ |
| Tamil Nadu | 1.16 (+) | 0.091 | 0.16 (+) | 0.533 |
| Uttar Pradesh | 22.12 (–) | $6.107 \times 10^{-15}$ | 2.57 (–) | 0.011 |
| West Bengal | 5.18 (–) | 0.0003 | 8.06 (–) | $6 \times 10^{-6}$ |

The third column of Fig. 5 showed the correlation of the ℝ(*t*) with *AH* for ten Indian states which were badly affected by the COVID-19. Most of the states showed a negative correlation between ℝ(*t*) and *AH*, which implied that the effective reproduction number decreased with an increase in absolute humidity. Absolute humidity may be playing a role to control the spread of the virus.

## 5 Discussion

There were plenty of studies that focused on the effect of humidity and temperature on virus transmission [26, 27, 28, 17, 29]. Our study is unique as we deal with time-dependent models and look for the impact of temperature and humidity on effective reproduction number which is the driving factor of a pandemic. Most of the earlier studies focused on the variation of infected individuals on temperature and humidity. Coronavirus spread has been influenced by a variety of parameters including personal hygiene, social distance, vaccination, etc. We assumed that these factors remained constant for these states throughout the study and that their impact on the result is minimal.

Earlier a mixed result was found between correlation of COVID-19 transmission and temperature. A positive correlation was found for several studies in New York [30] and Jakarta [31]. A negative correlation was observed for multiple worldwide studies [26, 27, 28, 17, 29, 32, 33] in California [25], Italy [34], Ghana [35], Japan [36], Spain [37, 38], and China [39, 40, 41, 42]. For the countries like Nigeria[43], Spain [44], Iran [45, 46], and in a worldwide study [47] no correlation was found. Global research of 166 nations (excluding China) found a negative correlation between temperature and COVID-19 cases, with a temperature increase of 1*°C* resulting in a 3.08% drop in cases [48]. Another worldwide study with 100 nations (up to 18 March 2020) with wide temperature variation from –33.9 to 34.3 *°C*, suggested a negative correlation between daily COVID-19 cases and temperature [49]. A significant negative correlation was reported earlier between daily COVID-19 cases and temperature (−10*°C* to 10*°C*) for provinces of China [41]. Several studies investigated the effect of humidity on the spread of the virus, but the results were inconclusive. There was no definite correlation between humidity and COVID-19 transmission in studies from Africa, Indonesia, Jakarta, New York, USA, and a global study of 100 nations [49, 30, 50, 31]. A weak positive correlation was found between average relative humidity and COVID-19 cases in Saudi Arabia [51]. A significant positive correlation between relative humidity and new COVID-19 cases was reported for the United States [52]. A negative correlation between the relative humidity and COVID-19 cases was reported for Spain [53], where transmission reduced by 3% for every 1% increase in humidity.

We used the most efficient time-dependent models SIRD and SEIRD to study the evolution of different time-dependent driving parameters of a pandemic like effective reproduction number and effective contact rate for different Indian states which were badly affected by COVID-19 for a long time scale of nearly eight months. During the time of our study, the infected cases were very high and we also focused on the most destructive second wave in India, which broke down all the health systems drastically. We also studied the evolution of time-dependent recovery rate and mortality rate. The mortality rate showed several spikes in the diagram which implied the dangerous impact of the virus on the population.

One of the main aspects of the study was to look for a correlation between one of the driving parameters of a pandemic the effective reproduction number with temperature and humidity using different time-dependent models. The correlation between the temperature and effective reproduction number was not conclusive. Few Indian states showed a positive correlation whereas few states showed a negative correlation. The positive correlation between the effective reproduction number and the average air temperature was prominent for the states West Bengal, Tamil Nadu, and Maharastra. A negative correlation was observed for the states Delhi and Rajasthan for both SIRD and SEIRD models. For the state of Madhya Pradesh, the correlation between the effective reproduction number and the average air temperature was negative for the SEIRD model, which is statistically significant (p-value<0.05).

The study of the correlation between the effective reproduction number and the humidity provided some important outcomes. We used both relative and absolute humidity to look for the correlation with effective reproduction number. All ten states showed a negative correlation between effective reproduction number and relative humidity with high statistical significance for the time-dependent SIRD model. For the state, Delhi the negative correlation significance was relatively low for the SIRD model which showed a positive correlation for the SEIRD model with higher statistical significance. The SEIRD model also showed a negative correlation between the effective reproduction number and relative humidity for all the states except Delhi.

The correlation between the absolute humidity and effective reproduction number showed a similar type of trend in correlation like relative humidity. All of the states except Tamil Nadu and Kerala showed a negative correlation between the absolute humidity and effective reproduction number for the SIRD model. The state Tamil Nadu showed a positive correlation with a high p-value, which was statistically less significant. For the SEIRD model in the state of Delhi, Kerala, and Tamil Nadu, the correlation between the effective reproduction number and absolute humidity was positive, the rest of the states showed a negative correlation. The p-value for the states Kerala and Tamil Nadu was high which implied less statistical significance.

## 6 Conclusion

We looked for the evolution of different driving parameters of a pandemic using time-dependent epidemiological models SIRD and SEIRD for different Indian states. We studied the correlation between effective reproduction number with average temperature, relative humidity, and absolute humidity of different Indian states. We found that most of the Indian states showed a negative correlation between effective reproduction number and relative and absolute humidity for time-dependent models SIRD and SEIRD, which implies that humidity may have an impact on the spread of the virus. Whereas, the correlation between temperature and effective reproduction number is not conclusive. There also exist several factors like personal hygiene, vaccination, social interaction, etc., which also played a crucial role in the spread of the virus. Environmental variables, particularly temperature and humidity, may play a role in epidemic spread, but their impact is marginal when compared to other social factors such as social interaction, governmental interventions, lock-downs, population density, herd immunity, personal hygiene, migration patterns, defense mechanisms and so on. Warm and moist environments appear to decrease the transmission of COVID-19. Temperature and humidity alone could not account for the majority of the variation in disease transmission. Therefore, countries should put more emphasis on healthcare policies and vaccination.

## Data Availability

We have used data from data.covid19india.org and https://github.com/CSSEGISandData/COVID-19.

## Declaration

### Ethics approval

Not applicable

### Consent to participate

Not applicable

### Avaliability of data and material

We have used data from data.covid19india.org/ and https://github.com/CSSEGISandData/COVID-19.

### Code availability

Not applicable

## Author contribution

Mr. Manik made data analysis and visualization of results. Mr. Mandal contributed to write the manuscript. Dr. Pal contributed to the conceptualization of the project and the manuscript writing. Mr. Patra and Mr Acharya took part in the manuscript writing and analyzing preliminary data.

## Footnotes

1 https://data.covid19india.org/

2 https://www.indiacensus.net/

## References

[1] P. Delhi, “Update on novel coronavirus: one positive case reported in kerala,” Ministry of Health and Family Welfare, vol. 1601095, 2020.

[2] Worldometer, “Coronavirus cases,” Worldometer, vol. 164, pp. 1–22, 2020.

[3] Worldometer, “Countries in the world by population (2021),” 2020.

[4] R. Ranjan, A. Sharma, and M. K. Verma, “Characterization of the second wave of covid-19 in india,” medRxiv, 2021. [Online]. Available: https://doi.org/10.1101/2021.04.17.21255665

[5] “Guidelines for international arrivals, ministry of health and family welfare, government of india, 17 february 2021,” 2021.

[6] “Genome sequencing by insacog shows variants of concern and a novel variant in india, ministry of health and family welfare, government of india, release id: 1707177,” 2021.

[7] W. H. Organization et al., “Covid-19 weekly epidemiological update, 11 may 2021,” World Health Organization, 2021.

[8] J. Shaman and M. Kohn, “Absolute humidity modulates influenza survival, transmission, and seasonality,” Proceedings of the National Academy of Sciences of the United States of America, vol. 106, pp. 3243–8, 03 2009. [Online]. Available: https://doi.org/10.1073/pnas.0806852106

[9] L. Poole, “Seasonal influences on the spread of sars-cov-2 (covid19), causality, and forecastabililty (3-15-2020),” 2020.

[10] Q. Bukhari and Y. Jameel, “Will coronavirus pandemic diminish by summer?” SSRN Electronic Journal, 01 2020. [Online]. Available: https://doi.org/10.2139/ssrn.3556998

[11] K.-H. Chan, J. S. Peiris, S. Lam, L. Poon, Y. ky, and W. H. Seto, “The effects of temperature and relative humidity on the viability of the sars coronavirus,” Advances in virology, vol. 2011, p. 734690, 10 2011. [Online]. Available: 10.1155/2011/734690

[12] S. Manik, S. Pal, M. Mandal, and M. Hazra, “Effect of 2021 assembly election in india on covid-19 transmission,” Nonlinear Dynamics, 2021. [Online]. Available: https://doi.org/10.1007/s11071-021-07041-7

[13] R. Gupta, G. Pandey, P. Chaudhary, and S. Pal, “Seir and regression model based covid-19 outbreak predictions in india,” medRxiv, 2020. [Online]. Available: https://doi.org/10.1101/2020.04.01.20049825

[14] E. L. Piccolomini and F. Zama, “Preliminary analysis of covid-19 spread in italy with an adaptive seird model,” 2020.

[15] L. Pribylova and V. Hajnova, “Seiar model with asymptomatic cohort and consequences to efficiency of quarantine government measures in covid-19 epidemic,” 2020.

[16] B. M. Ndiaye, L. Tendeng, and D. Seck, “Comparative prediction of confirmed cases with covid-19 pandemic by machine learning, deterministic and stochastic sir models,” 2020.

[17] A. Notari, “Temperature dependence of covid-19 transmission,” Science of The Total Environment, vol. 763, p. 144390, 2021. [Online]. Available: https://doi.org/10.1016/j.scitotenv.2020.144390

[18] S. Bhattacharjee, “Statistical investigation of relationship between spread of coronavirus disease (covid-19) and environmental factors based on study of four mostly affected places of china and five mostly affected places of italy,” 2020.

[19] S. G. Baum, “Covid-19 incubation period: An update,” NEJM Journal Watch, 2020.

[20] I. I. Bogoch, A. Watts, A. Thomas-Bachli, C. Huber, M. U. G. Kraemer, and K. Khan, “Pneumonia of unknown aetiology in Wuhan, China: potential for international spread via commercial air travel,” Journal of Travel Medicine, vol. 27, no. 2, 01 2020. [Online]. Available: https://doi.org/10.1093/jtm/taaa008

[21] S. Zhao, Q. Lin, J. Ran, S. S. Musa, G. Yang, W. Wang, Y. Lou, D. Gao, L. Yang, D. He, and M. H. Wang, “Preliminary estimation of the basic reproduction number of novel coronavirus (2019-ncov) in china, from 2019 to 2020: A data-driven analysis in the early phase of the outbreak,” International Journal of Infectious Diseases, vol. 92, pp. 214–217, 2020. [Online]. Available: https://doi.org/10.1016/j.ijid.2020.01.050

[22] M. Li, An Introduction to Mathematical Modeling of Infectious Diseases, 01 2018. [Online]. Available: https://doi.org/10.1007/978-3-319-72122-4

[23] E. Dong, H. Du, and L. Gardner, “An interactive web-based dashboard to track covid-19 in real time,” The Lancet Infectious Diseases, vol. 20, 02 2020. [Online]. Available: 10.1016/S1473-3099(20)30120-1

[24] A. Auler, F. Cássaro, V. da Silva, and L. Pires, “Evidence that high temperatures and intermediate relative humidity might favor the spread of covid-19 in tropical climate: A case study for the most affected brazilian cities,” Science of The Total Environment, vol. 729, p. 139090, 2020. [Online]. Available: https://doi.org/10.1016/j.scitotenv.2020.139090

[25] S. Gupta, G. S. Raghuwanshi, and A. Chanda, “Effect of weather on covid-19 spread in the us: A prediction model for india in 2020,” Science of The Total Environment, vol. 728, p. 138860, 2020. [Online]. Available: https://doi.org/10.1016/j.scitotenv.2020.138860

[26] M. Arumugam, B. Menon, and S. Narayan, “Ambient temperature and covid-19 incidence rates: An opportunity for intervention?” 04 2020.

[27] J. Chiyomaru and K. Takemoto, “Global covid-19 transmission rate is influenced by precipitation seasonality and the speed of climate temperature warming,” 04 2020. [Online]. Available: https://doi.org/10.1101/2020.04.10.20060459

[28] B. Pirouz, A. Golmohammadi, H. Masouleh, G. Violini, and B. Pirouz, “Relationship between average daily temperature and average cumulative daily rate of confirmed cases of covid-19,” 04 2020. [Online]. Available: https://doi.org/10.1101/2020.04.10.20059337

[29] M. M. Sajadi, P. Habibzadeh, A. Vintzileos, S. Shokouhi, F. Miralles-Wilhelm, and A. Amoroso, “Temperature, Humidity, and Latitude Analysis to Estimate Potential Spread and Seasonality of Coronavirus Disease 2019 (COVID-19),” JAMA Network Open, vol. 3, no. 6, pp. e2 011 834–e2 011 834, 06 2020. [Online]. Available: https://doi.org/10.1001/jamanetworkopen.2020.11834

[30] M. F. Bashir, B. Ma Bilal, B. Komal, M. Bashir, D. Tan, and M. Bashir, “Correlation between climate indicators and covid-19 pandemic in new york, usa,” Science of The Total Environment, vol. 728, p. 138835, 04 2020. [Online]. Available: https://doi.org/10.1016/j.scitotenv.2020.138835

[31] R. Tosepu, J. Gunawan, D. S. Effendy, L. O. A. I. Ahmad, H. Lestari, H. Bahar, and P. Asfian, “Correlation between weather and covid-19 pandemic in jakarta, indonesia,” Science of The Total Environment, vol. 725, p. 138436, 2020. [Online]. Available: https://doi.org/10.1016/j.scitotenv.2020.138436

[32] Y. Wu, W. Jing, J. Liu, Q. Ma, J. Yuan, Y. Wang, M. Du, and M. Liu, “Effects of temperature and humidity on the daily new cases and new deaths of covid-19 in 166 countries,” Science of The Total Environment, vol. 729, p. 139051, 2020. [Online]. Available: https://doi.org/10.1016/j.scitotenv.2020.139051

[33] J. Xie and Y. Zhu, “Association between ambient temperature and covid-19 infection in 122 cities from china,” Science of The Total Environment, vol. 724, p. 138201, 2020. [Online]. Available: https://doi.org/10.1016/j.scitotenv.2020.138201

[34] G. Livadiotis, “Statistical analysis of the impact of environmental temperature on the exponential growth rate of cases infected by covid-19,” PLOS ONE, 2020. [Online]. Available: https://doi.org/10.1371/journal.pone.0233875

[35] W. A. Iddrisu, P. Appiahene, and J. A. Kessie, “Effects of weather and policy intervention on COVID-19 infection in Ghana,” arXiv e-prints, p. 2005.00106, 2020.

[36] I. Ujiie, S. Tsuzuki, and N. Ohmagari, “Effect of temperature on the infectivity of covid-19,” International Journal of Infectious Diseases, vol. 95, 04 2020. [Online]. Available: https://doi.org/10.1016/j.ijid.2020.04.068

[37] A. Abdollahi and M. Rahbaralam, “Effect of temperature on the transmission of covid-19: A machine learning case study in spain,” medRxiv, 2020. [Online]. Available: https://doi.org/10.1101/2020.05.01.20087759

[38] Tobias and T. Molina, “Is temperature reducing the transmission of covid-19 ?” Environmental Research, vol. 186, p. 109553, 04 2020. [Online]. Available: https://doi.org/10.1016/j.envres.2020.109553

[39] A. Oliveiros, L. Caramelo, N. C. Ferreira, and F. Caramelo, “Role of temperature and humidity in the modulation of the doubling time of covid-19 cases,” medRxiv, 2020. [Online]. Available: https://doi.org/10.1101/2020.03.05.20031872

[40] H. Qi, S. Xiao, R. Shi, M. Ward, Y. Chen, W. Tu, Q. Su, W. Wang, X. Wang, and Z. Zhang, “Covid-19 transmission in mainland china is associated with temperature and humidity: A time-series analysis,” Science of The Total Environment, vol. 728, p. 138778, 04 2020. [Online]. Available: https://doi.org/10.1016/j.scitotenv.2020.138778

[41] P. Shi, Y. Dong, H. Yan, C. Zhao, X. Li, W. Liu, M. He, S. Tang, and S. Xi, “Impact of temperature on the dynamics of the covid-19 outbreak in china,” Science of The Total Environment, vol. 728, p. 138890, 2020. [Online]. Available: https://doi.org/10.1016/j.scitotenv.2020.138890

[42] A. Sil and V. N. Kumar, “Does weather affect the growth rate of covid-19, a study to comprehend transmission dynamics on human health,” Journal of Safety Science and Resilience, 2020. [Online]. Available: https://doi.org/10.1016/j.jnlssr.2020.06.004

[43] I. Taiwo and A. Fashola, “Covid-19 spread and average temperature distribution in nigeria,” SSRN Electronic Journal, 05 2020. [Online]. Available: https://dx.doi.org/10.2139/ssrn.3585374

[44] A. Briz-Redon and A. Serrano-Aroca, “A spatio-temporal analysis for exploring the effect of temperature on covid-19 early evolution in spain,” Science of The Total Environment, vol. 728, p. 138811, 04 2020. [Online]. Available: https://doi.org/10.1016/j.scitotenv.2020.138811

[45] I. Ahmadi, A. Sharifi, S. Dorosti, S. Ghoushchi, and N. Ghanbari, “Investigation of effective climatology parameters on covid-19 outbreak in iran,” Science of The Total Environment, vol. 729, p. 138705, 04 2020. [Online]. Available: https://doi.org/10.1016/j.scitotenv.2020.138705

[46] I. Jahangiri, M. Jahangiri, and M. Najafgholipour, “The sensitivity and specificity analyses of ambient temperature and population size on the transmission rate of the novel coronavirus (covid-19) in different provinces of iran,” Science of The Total Environment, vol. 728, p. 138872, 04 2020. [Online]. Available: https://doi.org/10.1016/j.scitotenv.2020.138872

[47] T. Jamil, I. Alam, T. Gojobori, and C. Duarte, “No evidence for temperature-dependence of the covid-19 epidemic,” Frontiers in Public Health, vol. 8, 08 2020. [Online]. Available: https://doi.org/10.3389/fpubh.2020.00436

[48] W. Yu, W. Jing, J. Liu, Q. Ma, J. Yuan, Y. Wang, M. Du, and M. Liu, “Effects of temperature and humidity on the daily new cases and new deaths of covid-19 in 166 countries,” Science of The Total Environment, vol. 729, p. 139051, 04 2020. [Online]. Available: https://doi.org/10.1016/j.scitotenv.2020.139051

[49] A. Meyer, R. Sadler, C. Faverjon, A. R. Cameron, and M. Bannister-Tyrrell, “Evidence that higher temperatures are associated with a marginally lower incidence of covid-19 cases,” Frontiers in Public Health, vol. 8, p. 367, 2020. [Online]. Available: https://doi.org/10.3389/fpubh.2020.00367

[50] I. A. Adekunle, S. A. Tella, K. O. Oyesiku, and I. O. Oseni, “Spatio-temporal analysis of meteorological factors in abating the spread of covid-19 in africa,” Heliyon, vol. 6, no. 8, p. e04749, 2020. [Online]. Available: https://doi.org/10.1016/j.heliyon.2020.e04749

[51] M. Alkhowailed, A. Shariq, F. Alqossayir, O. A. Alzahrani, Z. Rasheed, and W. Al Abdulmonem, “Impact of meteorological parameters on covid-19 pandemic: A comprehensive study from saudi arabia,” Informatics in Medicine Unlocked, vol. 20, p. 100418, 2020. [Online]. Available: https://doi.org/10.1016/j.imu.2020.100418

[52] L.-C. Chien and L.-W. A. Chen, “Meteorological impacts on the incidence of covid-19 in the u.s,” Stochastic Environmental Research and Risk Assessment, vol. 34, 07 2020. [Online]. Available: https://doi.org/10.1007/s00477-020-01835-8

[53] A. Paez, F. A. Lopez, T. Menezes, R. Cavalcanti, and M. G. d. R. Pitta, “A spatio-temporal analysis of the environmental correlates of covid-19 incidence in spain,” Geographical Analysis, vol. 53, no. 3, pp. 397–421, 2021. [Online]. Available: https://doi.org/10.1111/gean.12241

